# Estimates of SARS-CoV-2 infections and population immunity after the COVID-19 pandemic in Austria: Analysis of national wastewater data

**DOI:** 10.1101/2024.11.20.24317646

**Authors:** Uwe Riedmann, Alena Chalupka, Lukas Richter, Martin Sprenger, Wolfgang Rauch, Hannes Schenk, Robert Krause, Peter Willeit, Herbert Oberacher, Tracy Beth Høeg, John PA Ioannidis, Stefan Pilz

## Abstract

**Background:** Post-pandemic surveillance data on COVID-19 infections may help inform future public health policies regarding SARS-CoV-2 testing, vaccinations or other COVID-19 measures. We estimate the total SARS-CoV-2 infections in Austria after the end of the pandemic (May 5, 2023, per WHO) up to May 2024 from wastewater data. Those estimates are used in an agent-based model (ABM) to estimate average national levels of SARS-CoV-2 infection protection (IP) and COVID-19 death protection (DP).

**Methods:** We use a previously published model estimating total infections in Austria from wastewater data and extrapolate the approach up to May 2024. Utilizing those estimates in an ABM, we estimate daily national average IP and DP. These estimates are based on waning immunity estimates of previous literature and incorporate documented vaccinations.

**Findings:** We estimate approximately 3·2 million infections between May 6, 2023, and May 23, 2024, with a total of 17·8 million infections following May 12, 2020. The ABM estimates that 95% of people in Austria were infected with SARS-CoV-2 at least once. It also shows very high levels of national average DP a year after the end of the pandemic. National IP remained relatively low after the onset of Omicron.

**Interpretation:** The estimated high number of SARS-CoV-2 infections since the end of the COVID-19 pandemic in Austria has kept the national average DP very high. These findings should be considered for public health decisions on SARS-CoV-2 testing practices and vaccine booster administrations.

**Funding:** Austrian Science Fund (FWF) KLI 1188.

## INTRODUCTION

Immune conferring events by SARS-CoV-2 vaccinations and infections were critical to mitigate the COVID-19 disease burden resulting in significantly declining infection fatality rates (IFR) and the declaration by the WHO of the end of the COVID-19 pandemic by May 5, 2023. Thereafter, active national surveillance data based on SARS-CoV-2 testing and tracking of severe and fatal COVID-19 cases are largely missing. Post-pandemic surveillance data on COVID-19, however, may be required to inform future public health policies regarding SARS-CoV-2 testing, vaccinations or other COVID-19 measures. In particular estimates of the potential COVID-19 disease burden such as the IFR are important to guide us on how to balance the risks and benefits of any recommendation regarding COVID-19.

Estimating the number of SARS-CoV-2 infections based on wastewater data in the post-pandemic phase provides a measure on the extent of immune conferring events and thus on the immunological protection against COVID-19 in the general population.^1,2^ Immunological protection conferred after SARS-CoV-2 infections, termed natural immunity, may be superior to vaccine induced immunity as it wanes slower regarding protection against infection.^3^ Importantly, immunity by SARS-CoV-2 vaccination, infection and a combination thereof, termed hybrid immunity, all provide significant long-term protection against severe and fatal COVID-19 that shows little evidence of waning compared to the relatively short-lived protection against infections.^3–6^ In line with this, SARS-CoV-2 infection rates were still relatively high towards the end of the COVID-19 pandemic, boosting immunity in the general population, whereas IFR continuously declined. How immunity to SARS-CoV-2 and IFR further evolved in the post-pandemic phase is, however, largely unknown.

In this study we used a previously published model on national wastewater data to estimate the number of all SARS-CoV-2 infections that occurred in Austria after the declared end of the COVID-19 pandemic from May 6, 2023 to May 23, 2024.^1^ In addition, we estimate the nationwide average protection against COVID-19 death (DP) and protection against SARS-CoV-2 infections (IP) by using an agent-based model (ABM) as an extension of an SIR (susceptible - infectious – recovered) model that estimates waning immunity according to literature-based data and incorporates the estimated total infections and documented vaccinations.^3–5,7–14^

## METHODS

### Design and analysis

We conducted a retrospective estimation of total SARS-CoV-2 infections in the entire population of Austria from May 12, 2020 to May 23, 2024, based on wastewater monitoring data.^1,2,15^ Additionally, an ABM was constructed to estimate the temporal changes of the nationwide level of immunization in form of DP and IP. DP and IP reflect how much lower the probability of death and infection, respectively, is when compared to immunity-naïve individuals. This is akin to how vaccine effectiveness is often calculated (1-Hazard Ratio).^16,17^ The model was based on individual levels of DP and IP after a previous infection, previous vaccination (one, two and three or more doses), a hybrid immunization (at least one infection and one vaccination) and their respective rates of waning, retrieved from the literature.^3–5,7–14^ We used the 4-month moving average DP and regressed them onto 4-month moving average IFR estimates after April 2020.^18^ We also similarly regressed estimates from January 2022 onward, coinciding with Omicron dominance. The IFRs were estimated by dividing all COVID-19 deaths that occurred within a 4 month range around a date, by all first positive tests of an infection that occurred in that same period. To account for time lag between infection and death, we handled deaths as if they occurred on the first day of the last recorded SARS-CoV-2 infection which led to the death. Subsequent positive tests were counted as new infections if they occurred at least 90 days later. For the estimates we used 30-day COVID-19 mortality.^19^

The study was approved by the ethics committee at the Medical University of Graz (no. 33-144 ex 20/21). Analyses were pre-specified and agreed among authors before any data were analysed. The statistical analyses and simulations were conducted using R (version 4.4.1).^20^

### Study data

Wastewater data from the Austrian SARS-CoV-2 wastewater monitoring initiative was provided by the Austrian Federal Ministry of Social Affairs, Health, Care and Consumer Protection for the period from November 1, 2022, to May 31, 2024. Estimates of daily active infections between May 2020 and December 2022 were provided by a previous publication.^1^ Data on nationwide daily vaccinations and number of doses was publicly available for the time period between December 27, 2020 and January 1, 2024.^21^ As in previous publications, documented SARS-CoV-2 infections and COVID-19 deaths were provided by the Austrian Agency for Health and Food Safety (German: Österreichische Agentur für Gesundheit und Ernährungssicherheit; AGES) and acquired through the Austrian epidemiological reporting system (German: Epidemiologisches Meldesystem; EMS).^16,22^

### Models

#### Infection estimation from wastewater data

For the estimation of total SARS-CoV-2 infections, we applied a previously published approach, that estimated infections in Austria based on wastewater data from May 2020 to December 2022.^1^ The model showed high overlap with two different approaches estimating total infections based on IFRs and test positivity (testing rate and reported cases).^1^ Active infection estimates from the original publications were available between April 30, 2020, and December 17, 2022, and the model was extrapolated up to May 31, 2024, using the respective wastewater data. Usage of backpropagation for estimation of daily new infections (see Supplements) led to the earliest estimates of new daily infections on May 12, 2020, and latest on May 23, 2024.

A description on the wastewater monitoring data as well as the pre-processing and normalization methodology is presented in the Supplements, and more detailed in previous publications.^1,2,15,23,24^ In short, the data was filtered, outlier corrected, averaged and interpolated before the model estimation. The model is based on a parameter estimate that represents a combined shedding and loss factor (how many gen copies can actually be identified per infection) which the original paper estimates for multiple timeframes.^1^ We adopted the last used parameter estimate (from May 2022 onward) to extrapolate the analysis up to May 31, 2024. We conducted preliminary analyses to recalibrate the parameter by varying degrees and incorporated these adjustments into the agent-based model (ABM) that accounts for reinfection risk. Our findings indicated that reducing the parameter estimates by 25% (equivalent to increasing the estimated daily infections by 25%) produced population outcomes aligned with later seroprevalence studies (Table S3).^25^ Consequently, we adopted these recalibrated estimates for the main analysis.

We correlated recorded infections with the estimated total infections between December 17, 2022, and June 30, 2023, as an indication of how well documented fluctuations in infections are represented in the estimates. This marks the period in which the previous study did not estimate total infections, but infections were still officially documented. For a detailed description of the model see Supplementary Methods and the original publication.^1^

#### Agent-based model

We constructed an ABM to simulate the immunity level (DP and IP) of the general population of Austria between May 12, 2020, and May 23, 2024. It was conceptualized as an extended SIR model where the individual state of every agent is tracked, and previously estimated infections are probabilistically distributed based on the individual IP levels (individual susceptibility = 1-IP) (Figure 1). We additionally included the option of vaccination to move from S to R. The state tracks number of previous infections, number of previous vaccinations, days since last infection and days since last vaccination for every agent in R (Figure 1). From this state we categorized every agent into one of five compartments (natural immunity; vaccinated once; twice; three or more times; and hybrid immunity). Agents in these categories have a non-zero IP which wanes over time, the rate of which is based on published data estimates of IP waning in the respective category.^3–5,7–14^ Agents in S have an IP of zero.

**Figure 1:**
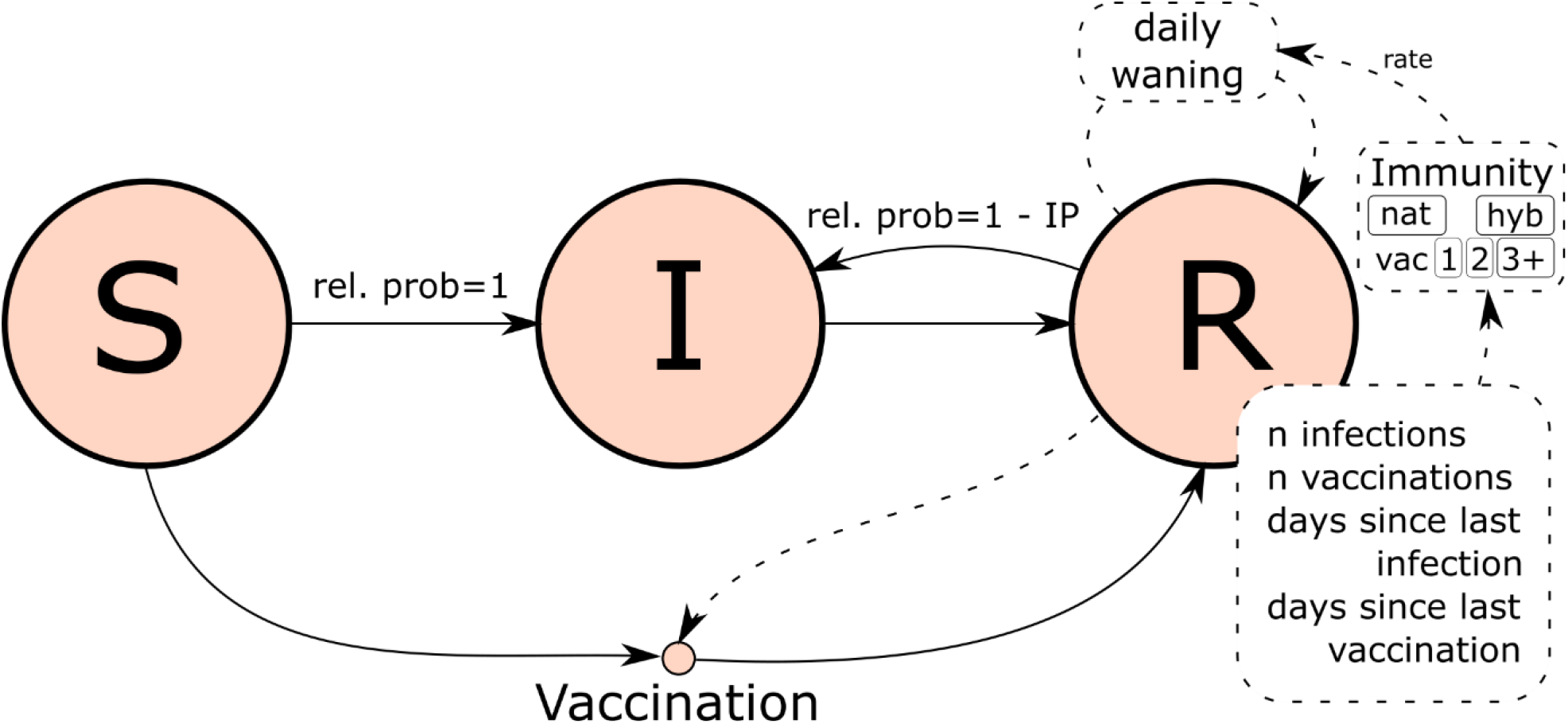
IP and DP estimation model concept. Agents can be in state S, I or R. Recovered agents save state information about previous events to group them into natural immunity, vaccinated with 1, 2 or 3+ doses and hybrid immunity. Group specific waning rates and information on days since last infection are then used to calculate the daily IP and DP.

Infected agents are in I for 14 days before moving to their respective R compartment. In this time they cannot be infected or vaccinated. We additionally tracked the daily individual DP, also based on published data estimates of DP specific waning.^3–5,10–13^ By averaging the individual IP and DP respectively, excluding currently infected individuals, we calculated the level of daily average national IP and DP. Vaccinations were lagged 14 days to account for the time needed to become effective. We set the population to 9,020,000.^1^

Waning protection was implemented using a Gompertz function.^4^ To estimate the parameter for the function based on waning estimates, we fitted the Gompertz curve to the waning estimates using non-linear least squares.^4^

We retrieved individual estimates of waning immunity for first, second and three or more vaccinations, as well as estimates for previous infections and hybrid immunity from previous publications.^3–5,7–14^ For a description of waning immunity estimates including sources and values, see Supplementary Methods (Table S1 and S2, Figure S1). If hybrid immunity occurred, or a subject with hybrid immunity had an additional event, protection against reinfection and death were increased to the same level irrespective of number of infections, vaccination dose and event order.^9^ Studies show much lower IP in the Omicron period than before. So we researched separate estimates for pre-Omicron and Omicron periods and continuously transitioned waning estimates of individual IPs between them throughout a month in early 2022, at the onset of Omicron.^4^ There is no randomized data about the effectiveness about COVID-19 vaccines in the setting of the Omicron variant. Existing literature does not indicate that Omicron led to a decrease of protection from death, however the methodology used to assess for vaccine effectiveness against death may be inherently biased.^27,28^ For more details and a full mathematical description of the model please see the Supplementary Methods.

Our pre-analyses showed that variability of estimates decreased with lower simulation scaling (see Supplementary Methods; Figure S3). Thus, we decided to run the main simulation 15 times at a scale factor of 10. Scaling was performed by dividing daily estimated infections, vaccinations and the population by the scaling factor and rounding. The approach in this paper constitutes a conceptual extension to previously published investigations on gradual waning of immunity.^29–31^ We accounted for various sorts of bias like healthy vaccinee bias in the sensitivity analyses.^16,17,32–34^

#### Sensitivity analyses

We conducted sensitivity analyses on the parameters of waning in previously infected, one two or more times vaccinated and hybrid immunized individuals, by rerunning the simulation with waning DP 110%, 90% and 75% the magnitude of the original waning estimates. We also decrease IP estimates for previous infection IP only, infection plus hybrid IP and all three vaccination IP conditions to 75% to see the effects on infection distributions. We addressed the possibility of overestimating or underestimating the number of infections after the end of the pandemic by performing the main analysis with 125%, 110%, 90% and 75% of the estimated daily infections. To investigate the possibility of over and underestimating infections throughout the whole pandemic we also ran it with 125%, 110%, 90% and 75% of the estimated daily infections. We addressed healthy vaccinee bias by running the simulation with decreased DP for vaccinated individuals (including the one; two; and three or more vaccination categories), using 75% and 50% of the main simulation magnitude. Some literature indicates no waning of DP in previously infected and vaccinated.^4,5^ To address this possibility we ran the simulation with alternative estimates for infection and hybrid DP with no waning and estimates that were between these and the original estimates. Lastly, we ran the simulation without vaccinations. To decrease computational demand, we downscaled the sensitivity analyses by a factor of 200 while still running them 15 times each.

## RESULTS

### Infection estimation from wastewater data

The model estimated a total of approximately 3·2 million infections between May 6, 2023, and May 31, 2024. Between May 12, 2020, and May 23, 2024, 17·8 million total infections were estimated in Austria (Figure 2; see Table S3 for exact estimates used in ABM; also see Figure S4 and Supplementary results for confidence intervals). Including the 20,208,999 documented vaccinations, this totals close to 38 million immune conferring events (Figure 2). Extrapolated estimates for the period between December 17, 2022, and June 30, 2023, were correlated by r = 0.985 with seven day averaged documented infections (Figure S5)

**Figure 2:**
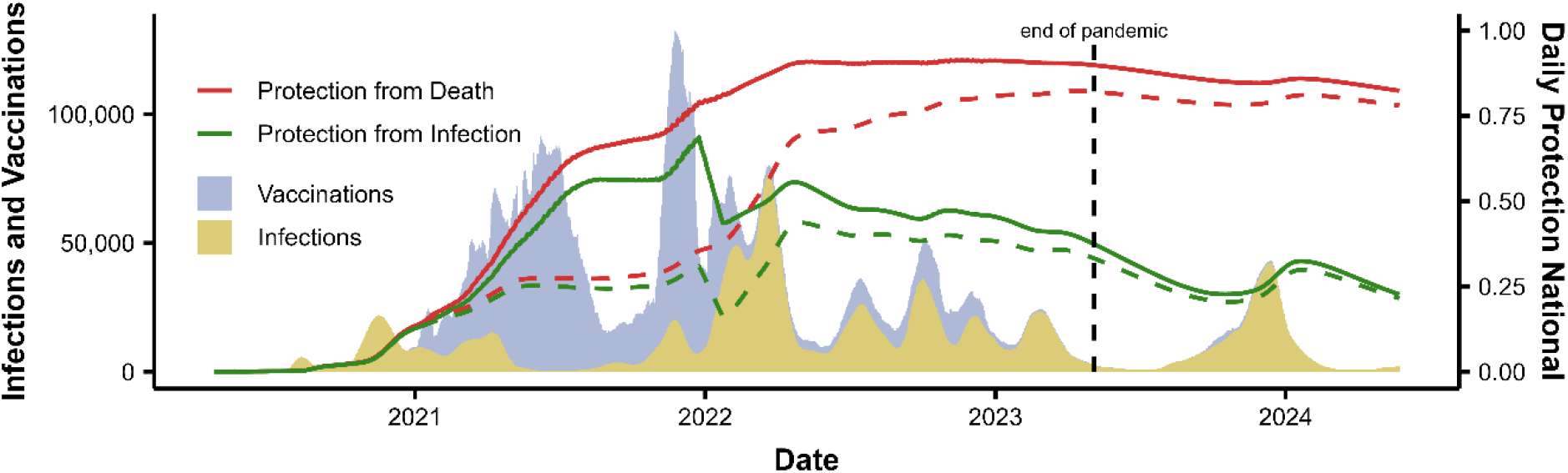
DP (red) and protection from infection (green). Dashed lines are simulated estimates of protection levels without vaccinations.

### Agent based model

The agent-based model shows the steady rise of the relative protection of the Austrian population throughout the pandemic, with ongoing high average DP a year after the pandemic’s end (Figure 2 and 3). On March 5, 2023, and March 23, 2024, the estimated national average DP was approximately 90% and 82% respectively. Thus, the protection is still at 92% of the level it was at, by the end of the pandemic (Table S3).

**Figure 3:**
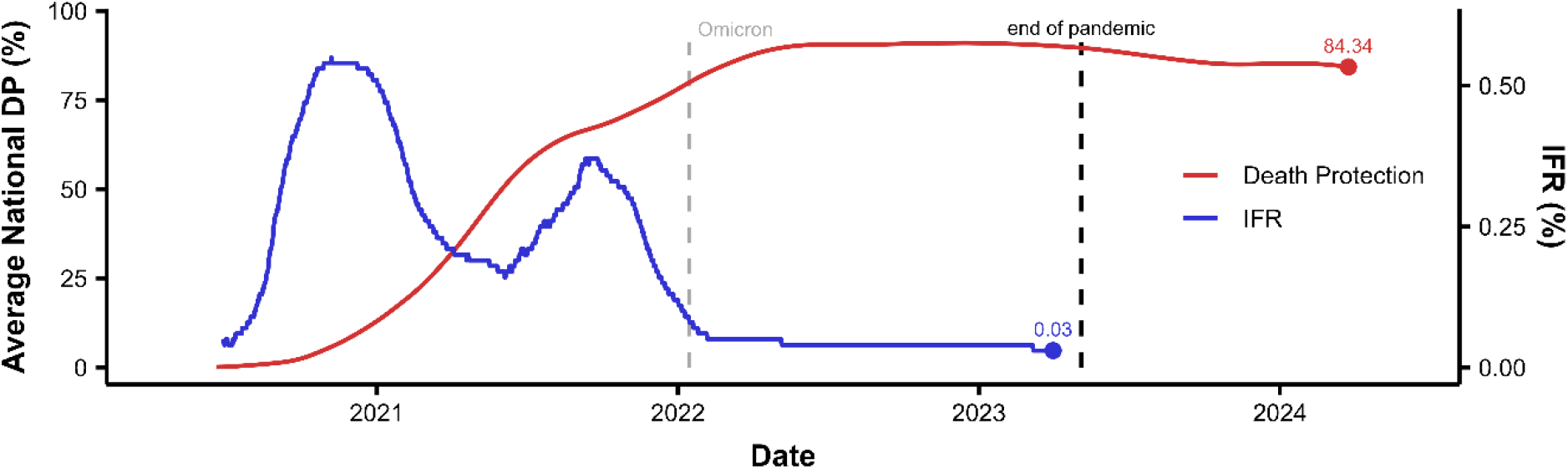
Comparison of average estimated protection from death (red) and IFR (blue). The IFR is based on the infection estimates and 30-day mortality data. DP and IFR represent 4-month moving averages. Note the difference in scales between IFR and DP.

Our model estimated that by May 23, 2024, a total of 99% of the Austrian population have had at least one immune conferring event, including 95% with at least one infection (Table S3). About 69% of the infected had multiple SARS-CoV-2 infections (Figure S8), and 47% of the population had their first infection before or without vaccination (Figure S9).

IFR decreased over the course of the pandemic, especially with the onset of Omicron (Figure 3). The last estimated IFR (April 1, 2023) was 0·03%. Four month rolling window DP predicted IFR estimates after June 29, 2020 significantly (R^2^ = 0·5246, F(1,1005) = 1104, p < ·001). Estimates after the onset of Omicron (January 2022) were also significantly predictive of IFR (R^2^ = 0·7681, F(1,454) = 1504, p < ·001) (Figure S10).

### Sensitivity analyses

Sensitivity analysis (Table S3 and Figure S11-S16) shows that the relative change in DP between May 5, 2023, and May 23, 2024, was DP never changed by more than 10%, irrespective of condition, with the biggest impact on this relative change being the potential overestimation and underestimation of infections and using alternative DPs that don’t wane (Table S3). Investigation of possible healthy vaccinee bias (downscaling protection against death provided by 1, 2 and 3+ vaccinations) showed little effect on national DP by May 2024 (Figure S14). Figure 2 shows the positive effect of vaccination in the population, as estimations excluding vaccinations show a much slower increase in DP across 2021 and 2022. The conditions with alternative DP estimates (slower or no waning) showed less to virtually no waning in national DP between May 2023 and May 2024 (Figure S15). For all results see Table S3 and Supplementary Results.

## DISCUSSION

We estimated that 3·2 million SARS-CoV-2 infections (about 35% of the population) occurred in Austria between May 6, 2023, and May 23, 2024. The ABM indicates high national DP from COVID-19 throughout this period even with the low number of recent vaccinations and no data on vaccination after January 2024. Sensitivity analyses showed that these findings are mostly robust to fluctuations in parameter choice. Reducing vaccination DP to account for a potential healthy vaccinee bias did not alter our findings that protection against death is high up to May 2024.

The findings suggest that Austria has maintained a high level of post pandemic protection against COVID-19 deaths, irrespective of vaccine boosters. The high number of estimated infections points to a high number of asymptomatic or mild cases of COVID-19, which is in line with very low IFR estimates at end of the pandemic. This may be due to a combination of very low IFR for the currently prevailing variants plus a high level of national DP. Our findings should be considered for public health policy regarding COVID-19 measures such as weighing the potential benefits and potential harms of SARS-CoV-2 testing.^35–38^

It further highlights the need to critically scrutinize the continued application of booster vaccines. As the number of individuals who have never been SARS-CoV-2 infected dwindles, and study data from Austria show that booster vaccinations may have no significant effect on the COVID-19 death risk for those who were previously infected, it is critical to cautiously balance the benefits with the risks and costs of further COVID-19 vaccine doses.^16^ Such considerations must take into account various adverse health consequences of SARS-CoV-2 infections beyond COVID-19 mortality but also the small but existing harms of vaccinations and the overall cost-effectiveness.

Results from the ABM further show that, even when accounting for possible infection overestimation, underestimation of waning and vaccine effectiveness overestimation, average national DP remains high. This may be in part due to the fact that since protection against infection wanes faster than protection against death, which leads to new infections in individuals before their DP can wane significantly. The very high number of asymptomatic or mild cases and the very low IFR support the hypothesis that the more time elapses since the beginning of the COVID-19 pandemic, the more SARS-CoV-2 resembles the endemic characteristics of the other human coronaviruses.^39,40^ This may raise the question, whether public health policy regarding SARS-CoV-2 should be similar as for the other endemic human coronaviruses.^41^

### Limitations

Extrapolation of total SARS-CoV-2 infection estimates was based on parameters value estimates from 2022, thus potentially limiting the validity of our estimates on the new daily infections from 2023 onward. We tried to address this in the pre-analyses and sensitivity analyses, by recalibrating parameter estimates and varying estimated infections (Figure S11). These estimates themselves are based on seroprevalence studies.^42,43^ Thus, potential errors may also be reflected in our ABM. While seroprevalence wanes and estimates late in the pandemic are potentially underestimations, the percentage of vaccinated individuals was unrepresentatively high leading to a potential overestimation. Consequently, individual seroprevalence estimates are not perfect indicators of population-level protection; however, the referenced seroprevalence estimates and our findings show significant alignment with estimates from various other countries..^44,45^

We also do not have total infection estimates before May, 12, 2020, even though this marks the period with the highest percentage of unidentified infections.^42^ As these should make up a relatively small part of overall infections, we do expect this to change our main conclusions, but it may mean the true rate of DP is higher than reported in this paper.

General limitations of infection estimation from wastewater data and differences in preprocessing regimes have been documented in previous publications ^46,47^

The ABM has multiple limitations based on available data and assumptions. For example, there are no data available on vaccination after January 1, 2024. As such the national protection against death may be underestimated thereafter, although vaccination rates were already exceedingly low by the end of 2023, e.g. only 62,172 (0·69%) people received a vaccine dose in November or December 2023 (Figure S2). We did not consider variations in effectiveness of different vaccine types.

Vaccine effectiveness estimates were, out of necessity, based on observational data, which is prone to multiple types of biases. Thus, there is inherent uncertainty about the amount and duration of IP and DP during the entire study period. We are not able to determine with our current methodology the exact extent to which vaccinations, prior infections and intrinsic virulence of the Omicron variant have independently affected the currently very low IFR (0·03%).

The model further doesn’t account for probabilities of infection and rates of waning moderated by age, comorbidities or other risk factors. As such, average national protection against death may be high, but some individuals may still have low levels of protection. It is unclear though whether vaccine boosters by late 2024 and thereafter might be effective even in this population sub-segment. The model also doesn’t account for changes in population based on births, deaths, immigration, emigration or aging. Additionally, simulation of protection trajectories without the application of vaccines do not consider any changes in the number of infected that such a scenario might have led to.

## Conclusion

This national investigation in Austria based on wastewater data estimates a high number of SARS-CoV-2 infections and thus a high level of immunological protection against COVID-19 deaths after the official end of the COVID-19 pandemic (i.e. May, 5, 2023). In light of previous studies on the potential harm of SARS-CoV-2 testing and studies showing booster vaccinations may not significantly increase protection from COVID-19 death in previously infected individuals, our findings support the ongoing recommendation against widespread SARS-CoV-2 testing and boosters for large parts of the general population.^16,35–38^ Further research on booster efficiency, average national protection levels and harm of testing as well as economical pressure of ongoing COVID-19 policies on the health care system is needed, to address possible up- and downsides of changes in those policies.

## Data Availability

The data that support the findings of this study are available upon request with approval
needed from the Austrian Federal Ministry of Social Affairs, Health, Care and Consumer
Protection and Austrian Agency for Health and Food Safety (German: Oesterreichische
Agentur fuer Gesundheit und Ernaehrungssicherheit; AGES) respectively.

## Contributors

UR and SP conceptualized the study with contributions of JPAI. UR and SP wrote the original draft of the manuscript. SP acquired funding. UR and HS wrote software. AC, LR, WR and HO curated data. UR performed the formal analyses and handled visualization. UR, HO and LR handled the validation. SP administered the project. All authors contributed to writing, reviewing, and editing the manuscript, and approved the final version before submission.

## Data sharing statement

The data that support the findings of this study are available upon request with approval needed from the Austrian Federal Ministry of Social Affairs, Health, Care and Consumer Protection and Austrian Agency for Health and Food Safety (German: Österreichische Agentur für Gesundheit und Ernährungssicherheit; AGES) respectively.

## Acknowledgements

The authors thank all persons and organizations involved in data collection. Funding: This work is funded by the Austrian Science Fund (FWF) KLI 1188. For this study, data from the National SARS-CoV-2 Wastewater Monitoring Program was used. This program is financed by the Austrian federal Ministry of Social Affairs, Health, Care and Consumer Protection who has the sole right of use for the data.

## Supplements

### Supplementary Methods

#### Wastewater Model

##### Data Description

The data from April 30, 2020 to December 17, 2022 were taken from Rauch et al., (2024).^1^ The extrapolation of this approach to March 30, 2024, is also based on this paper.

The dataset on wastewater data provided by the Austrian Federal Ministry of Social Affairs, Health, Care and Consumer Protection spanned the period from October 30, 2022, to May 31, 2024. It included 48 wastewater treatment plants (WWTP) from 2023 onward (24 before), with 7039 measurements total. These treatment plants cover approximately 58% of the Austrian population. Wastewater samples were collected twice a week.

Viral concentration is determined and processed as previously explained.^2^ Additionally, different hydrochemical parameters were used for characterizing the catchment population of the monitored wastewater treatment plants. For the chemical oxygen demand (COD) estimates, per capita equivalents were calculated using 120 g/d/person. Ammonia-nitrogen (NH4-N) estimates were calculated using 8.0 g/d/person. Total-nitrogen (Ntot) estimates were calculated using 11.0 g/d/person.^3^

##### Data Preprocessing

For a detailed description of preprocessing see previous publications.^1,2^

To compensate for inherent measurement noise, Rauch et al. (2021) suggest the approach to exclude outliers if the flow volume (Q) exceeds the 90 percentile of the long term recorded inflow data of a WWTP (needs at least a year of data points).^4^

Estimates are normalized based on population-size markers, to compensate for population fluctuations within a catchment area. Following Arabzadeh et al. (2021), we used NH4-N prioritised over COD and Ntot.^3^ We computed the daily weighted averages of viral load levels per federal state. The weights correspond to the design capacity of the respective WWTPs, prioritising large plants over smaller ones. The design capacity of each WWTP is a parameter, that serves as a weighting factor when computing the weighted average of multiple measurements in spatial aggregation. In principle, the preferred weighting factor is the exact catchment population. However, this information is currently unavailable to us.

This results in a scattered time-series from WBE measurements that are not gapless on a daily basis. To distribute the timeseries data equally both, up- and downsampling approaches are viable.^5^ Here we used upsampling to get daily estimates by linearly interpolating gaps before applying data smoothing. Lastly, data filtering techniques are applied to reduce the signal noise and provide a mechanism to obtain the underlying information of the signal.

##### Model Description

The measured virus load at the monitoring point is related to the population drained with the sewer system:

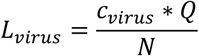

Where L_virus_ is the population normalized virus load in gene copies/**p**erson/**d**ay, Q is the flow volume in L/d, c_virus_ is the virus concentration in the sample in copies/L and N is the catchment population.

Under the assumption that each infected person is shedding a certain load of gene copies per day (L_shed_ in gene copies/P/d) into the sewer system and additionally introducing a general loss term f_loss_ we get:

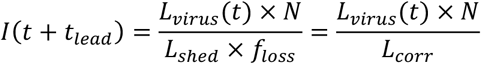

Where *I* is the number of infected individuals in the watershed, t_lead_ is the time lead and f_loss_ is a dimensionless loss factor.

Rauch chose t_lead_ = 7 in the original publication, based on cross correlation with documented infections.^1^ We found that the cross correlation of our estimates was highest with t_lead_ = 0, so we used that. Rauch and colleagues estimated different L_corr_ values for different timeframes. We chose to apply their estimate for the most recent timeframe (L_corr_ = 10^10.090). The population was set to *N* = 9.02 × 10^6^. As these values represent currently infected, we needed to apply a backcasting algorithm to estimate daily new infections.

##### Backcasting

The backcasting methodology used here is designed to estimate daily infections based on daily active cases. The key assumption is that an infection lasts 14 days on average.

We first smoothed the estimated undocumented daily infection counts via 14 days centred moving average. The core of the backcasting process involves iteratively refining the infection level estimates, which were initially based on the smoothed data. Potential estimation errors from variability in counting method was addressed by setting negative testing values to 0. Lastly, after refining the estimates, a secondary smoothing step was applied to the calculated infection values.

#### ABM Model

##### Model Description

Our novel ABM is an extension of the classic SIR framework that accounts for multiple immunity states, vaccination, and time-dependent waning immunity. It tracks recovery states for infections, different vaccination statuses and hybrid immunity, allowing for a nuanced representation of population-level immunity dynamics. The model uses a Gompertz function to simulate waning immunity based on time since last infection or vaccination, providing a novel approach to long-term epidemic modelling of national immunity levels.

*Note*: Conceptually this model is closer to the literature on extended SIRS model (Susceptible-Infected-Recovered-Susceptible) than on SIR frameworks.^6,7^ We decided to not introduce the possibility to move from R so S for multiple reasons: (1) mathematically the Gompertz function asymptotically approaches its upper limit. Thus, waning immunities would never truly be back to base level after an event. This may be addressed by setting a range at which immunity can be considered to have reached 0. (2) In the context of our data, it seems unreasonable that more than a few agents avoided infections for long enough for the waning to progress this far. (3) Most importantly, immunity levels against <u>infection</u> may wane enough to be comparable to agents in S, but waning of protection from death is estimated to be so slow that it won’t reach 0 in the timeframe of this study (more than 4 years). Thus, categorizing previous R grouped individuals into S, would be a misrepresentation of the underlying state. In other words: We do not think that grouping individuals with no IP but some level of DP into the category “Susceptible” is a sensible practice. This should be (re-)considered by researchers that plan to use this model on longer timescales, with lower infection rates, on different diseases or with different waning functions.

#### Model Structure

For each agent *i* at time *t*:

1. C_i_(t): State variable (0: Susceptible, 1: Infected, 2: Recovered)
2. I_i_(t): Infection status (0, 1+)
3. V_i_(t): Vaccination status (0, 1, 2, or 3+ doses)
4. T_i_(t): Time since last immunity-affecting event
5. P_R,i_(t): Protection level against infection
6. P_D,i_(t): Protection level against death
7. IM_i_(t): Immunity type
  a. no immunity (0)
  b. infection (1)
  c. vaccination doses
    i. one (2)
    ii. two (3)
    iii. Three or more (4)
  d. hybrid immunity (5)

#### Model Implementation

1. Initialize all N agents with all states = 0.
2. For each time step t:
  a. Update immunity levels P_R,i_(t) and P_D,i(_t) for all agents based on their immunity type IM_i_(t).
  b. Apply the **Infection Process** using infection data.
  c. Apply the **Vaccination Process** using vaccination data.
  d. Apply the **Recovery Process** for infected agents.
  e. Update immunity timer T_i_(t) for all agents.
  f. Collect population-level statistics, including distributions of immunity types and protection levels.

#### Immunity Dynamics

Gompertz-based waning immunity: Immunity wanes according to a Gompertz function, depending on immunity level i:

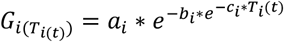

Protection from infection:

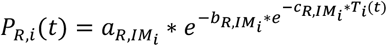

Protection from death:

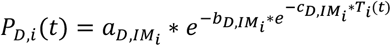

#### Immunity Type Update Rules

Upon infection:

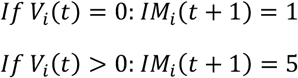

Upon vaccination:

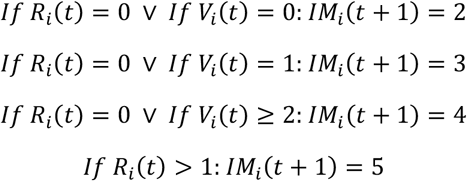

#### Infection Process

For each time step *t*:

1. Get the number of new infections I_new_(t) from estimate data.
2. Create a pool of potentially infectable individuals:

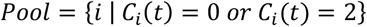
3. For each new infection (from 1 to I_new_(t)):
  a. Calculate selection probabilities for each individual i in Pool:

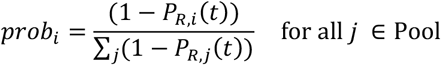
  b. Randomly select an individual based on these probabilities
  c. Update selected individual:

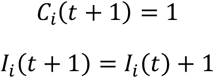
  d. Update the immunity type IM_i_ (t+1) according to Immunity Type Update Rules
  e. Reset immunity timer T_i_(t+1) = 0

#### Vaccination Process

For each time step t:

1. Get the number of new vaccinations for each dose 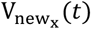, where x=1,2,3,4+ from vaccination data.
2. For each vaccination dose x from 1 to 4+:
  a. Create a pool of eligible individuals for each dose:

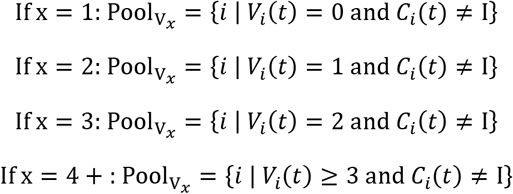
  b. For each new vaccination of dose x (from 1 to 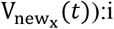:i. If 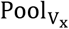 is not empty:
    i. Randomly select an individual i from 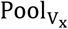
    ii. Update vaccination status: V_i_(t+1) = V_i_(t) + 1
    iii. Update immunity type IM_i_(t+1) according to Immunity Type Update Rules
    iv. Reset immunity timer: T_i_(t+1) = 0
    v. Update protection levels P_R,i_(t+1) and P_D,i_(t+1)
3. Update population-level vaccination statistics

#### Recovery Process

For each agent i with I_i_(t) = 1:

If the agent has been infected for 14 days:

1. Set C_i_(t+1) = 2
2. Update immunity type IM_i_(t+1) according to Immunity Type Update Rules
3. Reset immunity timer T_i_(t+1) = 0

#### Gompertz Curve Parameters

The parameter choice for the national protection from infection and death model was based on multiple previously published estimates.^8–18^ Estimation of Gomperz Curves Parameters were performed for pre-Omicron Era and Omicron Era. Protection was continuously changed from the pre-Omicron curve estimates to the Omicron curve estimates between December 25^th^ 2021 and January 25^st^ 2022. Different estimates for hybrid immunity showed consistently higher values than from a previous infection only.^12^ Interestingly, all of them showed relatively faster waning, this inevitably leads to a lower immunity level for hybrid immune than from previous infections. As waning estimates do are usually not performed or reliable on the time scale of multiple years, <u>we expect that the hybrid immunities are approaching the immunity of previously infected</u>. As such we set the lower bound for hybrid immunity at the respective waning immunity of the previous infected immunity (Figure S1).

##### Pre-Omicron Era

Table S1 show the waning estimates with their respective time points (in days) and the respective estimated start values and the reference from which this value was taken. Note that waning values <u>of protection against infection</u> were continuously reduced between December 25^th^ 2021 and January 25^st^ 2022 to Omicron period levels. This transition is not shown in the following figures as transition is infection time specific (Figure S1). The most reliable estimates on vaccination data in pre-Omicron Era were available for vaccine dose 2 (primary vaccination). As such we used point estimates for vaccination dose 1 and 3 to scale the dose 2 waning curve appropriately.

**Figure S1:**
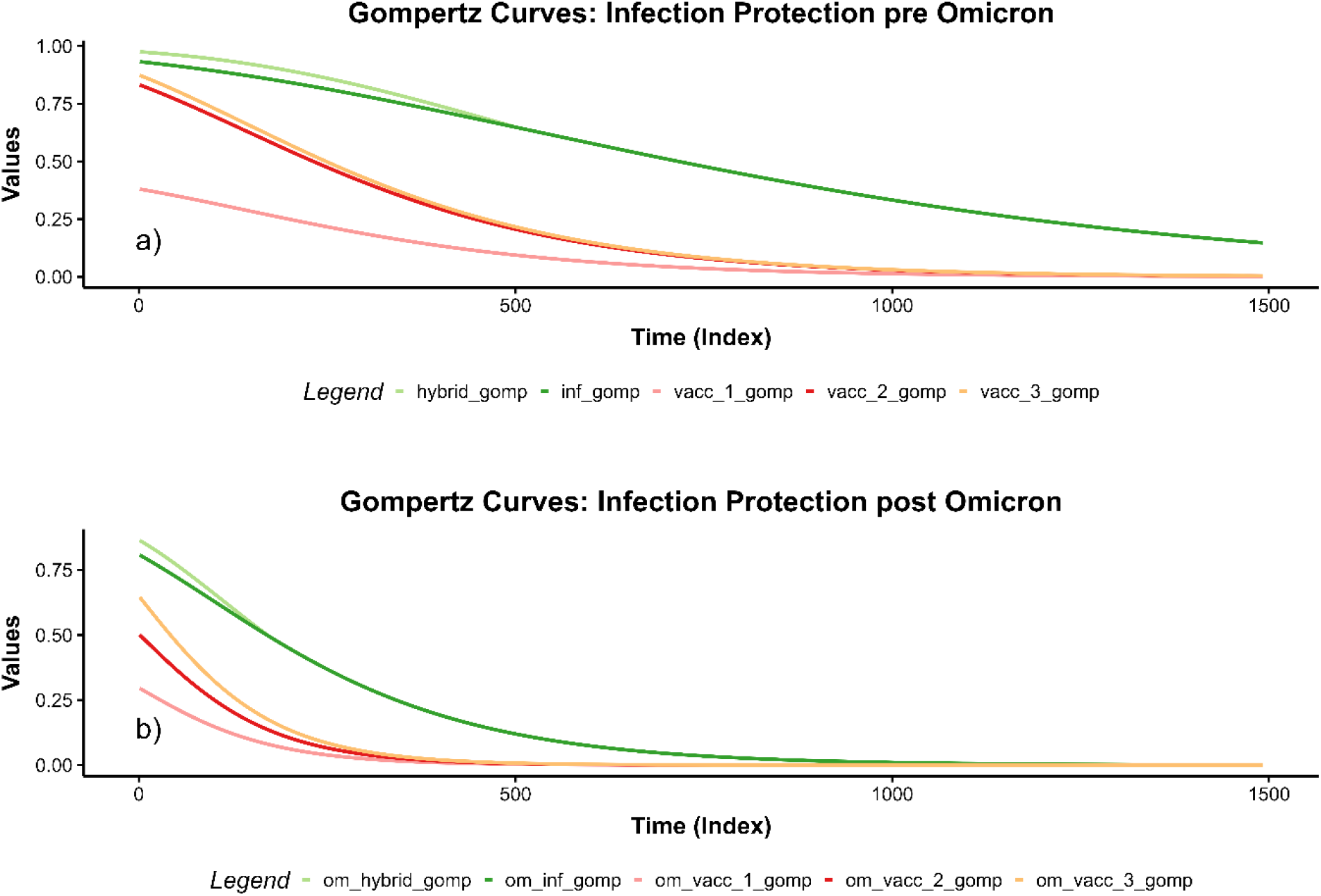
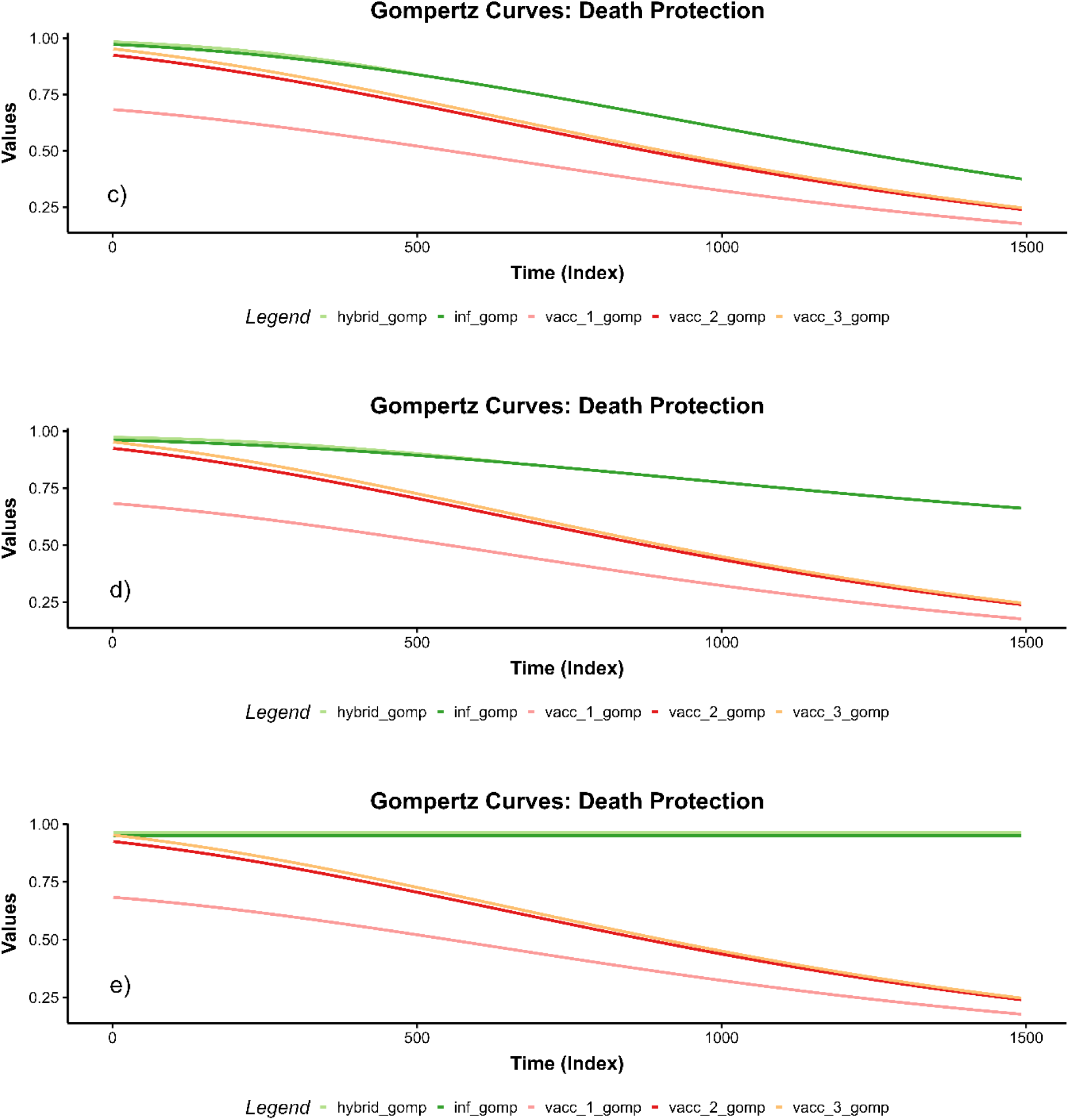
*Waning curves* for protection against infection pre-Omicron (a), protection against infection in Omicron (b), main protection against death (c), in-between alternative and main protection against death estimates (infection and hybrid immunity) (d) and alternative protection against death estimates (e). Hybrid immunity wanes faster than protection from a previous infection. We assumed that hybrid immunity wanes to the level of infection from a previous protection and then stays at that level.

##### Post-Omicron Era

The most reliable estimates on vaccination data in Omicron Era were available for vaccine dose 3 (booster vaccination). As such we used point estimates for vaccination dose 1 and 2 to scale the dose 3 waning curve appropriately (Figure S1b).

While some studies show small decreases in protection against death of previously infected in Omicron time periods, compared to pre-Omicron periods these estimates have two problems.^19^ First, the number of deaths in Omicron periods is so low, even for people without immunity, that a single death can already cause smaller protection estimations than in previous variants. Second, and more importantly, at this point of the pandemic, partially driven by the Omicron itself, the number of hidden infections is so high that even in test-negative designs the non-infected population is likely permeated by previously infected persons. Thus, indirectly decreasing the protection estimate. As most of the DP values for previously infected, are still extremely high, we thus concluded that the difference between pre-Omicron and Omicron DP estimates is likely to be an artefact. As such we did not adjust DP for previously infected, vaccinated or hybrid immune individuals at the onset of Omicron.

**Table S1:** Estimates of infection protection waning rates, references and reasoning of Gompertz functions.

|  | Estimated protection at day 1 (in percent) | Values (days: protection(%)) | Reference | Comment |
| --- | --- | --- | --- | --- |
| **pre-Omicron IP** |  |  |  |  |
| waning after infection | 93.24 | 14: 90.5, 487: 70, 669: 50 | Chemaitelly et al., 2022<sup>8</sup> | These are projected estimates |
| waning after 1 vaccination dose | 38.01 | | Chemaitelly et al., 2021<sup>9</sup> | We scaled the Gompertz curve estimates of 2 vaccination dose waning by the ratio between initial protection of 1 dose and 2 doses as given by Chemaitelly et al., 2021; $36.8\% / 80.5\% = 0.457$ |
| waning after 2 vaccination doses | 83.18 | 30: 80.5, 183: 54.6, 274: 45.9 | Menegale et al., 2023<sup>10</sup> |  |
| waning after 3 or more vaccination doses | 87.0 | | Braeye et al., 2023<sup>11</sup> | We scaled the Gompertz curve estimates of 2 vaccination dose waning by the ratio between initial protection of 3 doses and 2 doses as given by Braeye et al., 2023; $87\% / 83.18\% = 1.05$ |
| waning after hybrid immunity | 97.53 | 30: 96.58, 91: 96.1, 152: 90.25, 213: 89.17 | Goldberg et al., 2022<sup>12</sup> | Goldberg provides rate ratios with 2 vaccine doses as a reference value. We used the relative protection of 2 vaccine doses stated above to calculate these values from the rate ratios of hybrid immunity. |
| **Omicron IP** |  |  |  |  |
| waning after infection | 80.77 | 40: 77.3, 126: 53.0, 196: 45.5, 266: 37, 336: 37, 406: 37 | Covid-19 Forecasting Team, 2023<sup>13</sup> | Used values from Table S2 category: Protection against Omicron BA.2 reinfection |
| waning after 1 vaccination dose | 22.88 | | Chemaitelly et al., 2021<sup>9</sup> | We scaled the Gompertz curve estimates of 2 vaccination dose waning by the ratio between initial protection of 1 dose and 2 doses as given by Chemaitelly 2021 (pre Omicron ratio); $36.8\% / 80.5\% = 0.457$. We used the same ratio as pre Omicron as to our knowledge, there are no studies on effectiveness of partial vaccination against infection in the Omicron period. |
| waning after 2 vaccination doses | 50.06 | 30: 44.4, 182: 20.7, 274: 13.4 | Menegale et al., 2023<sup>10</sup> | These values present the estimates to calculate the value at day 1, not the values used for the actual Gompertz curves. We used the here estimated value to calculate a scaling from 3 vaccine doses. Menegale provides waning estimates for two doses, but these are likely biased to people that did not get a third vaccination dose. This is also indicated by the waning of IP which is much slower in the estimate of 2 doses than the estimate of 3 doses; $50.06\% / 64.6\% = .775$ |
| waning after 3 or more vaccination doses | 64.6 | 30: 55.4, 183: 36.0, 274: 28.9 | Menegale et al., 2023<sup>10</sup> |  |
| waning after hybrid immunity | 86.38 | 30: 80.1, 61: 74.8, 91: 68.6, 122: 61.6, 183: 46.5 | Bobrovitz et al., 2023<sup>14</sup> | We used the values for hybrid immunity (first booster vaccination) from Table 2. As these match more coherently with the waning after infection estimations, and are likely more reflective of omicron effects. |

Note: the inverse of the protection value (susceptibility) is used to estimate the Gompertz function

**Table S2:**
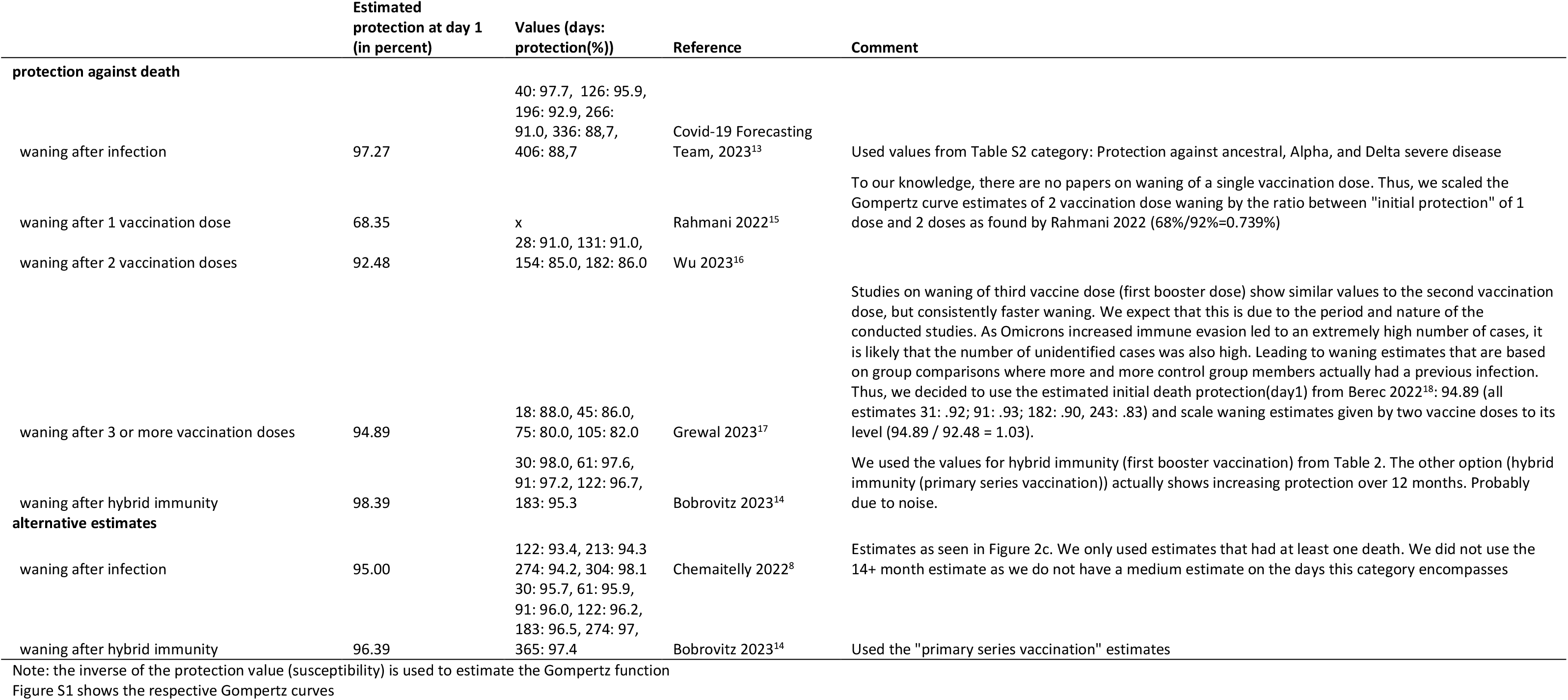
Estimates of death protection waning rates, references and reasoning of Gompertz functions.

#### Vaccination

**Figure S2:**
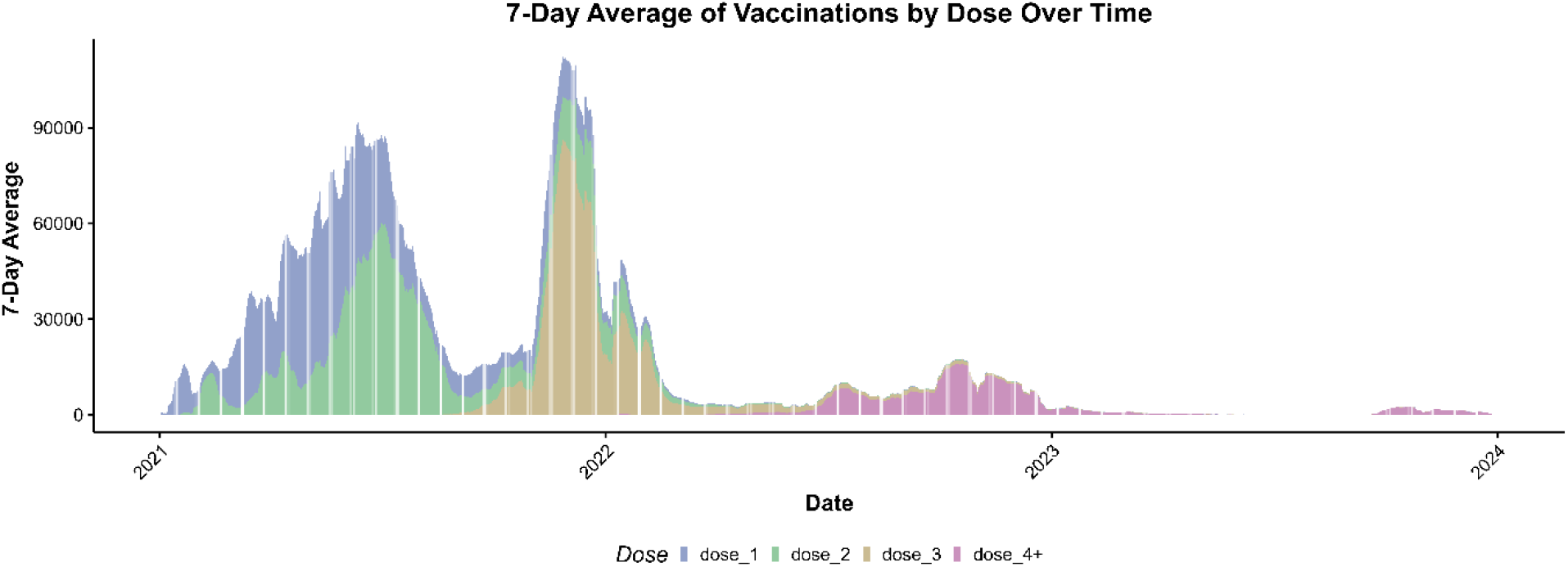
(7 Day Average) Vaccinations by Dose

#### Effects of Scaling on Variance between Runs

Early scaled analyses showed clearly that variations in IP and DP between multiple runs are small and keep decreasing with lower scaling. We calculated Root Mean Square Error (RMSE) to quantify the difference between multiple runs and based on these ever-decreasing variations (Figure S3),^20^ we concluded that using a low number of runs with a relatively smaller scaling is preferable (for computational viability) to higher scale, high run number averages.

**Figure S3:**
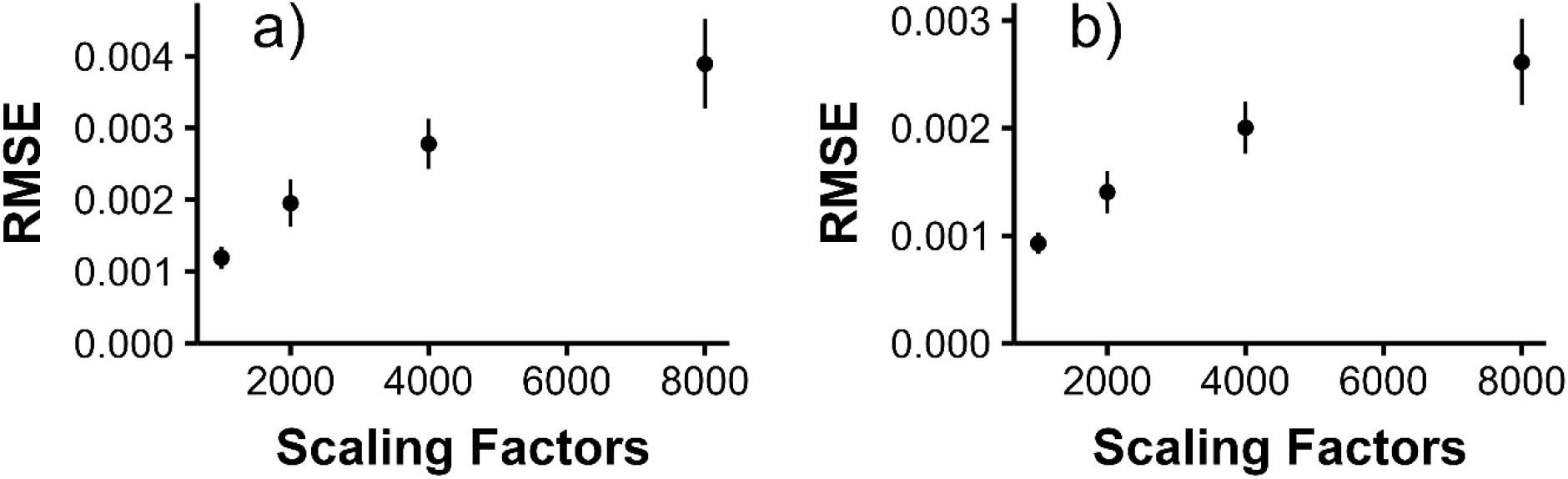
RMSE of DP (a) and IP (b) from 15 runs for different scaling factors.

#### Supplementary Results

##### Infection Estimation

As the original publication provided lower and upper bound estimates (5% and 95%) for the Lcorr parameter. As with the main analysis data, the estimates for these bounds prior to December 17, 2022 were acquired directly form Rauch et al.,(2024).^1^ After rescaling as explained in the main manuscript, we estimated a total of 18,701,194 and 17,165,579 infections for the 5% and 95% bounds respectively (Figure S4).

**Figure S4:**
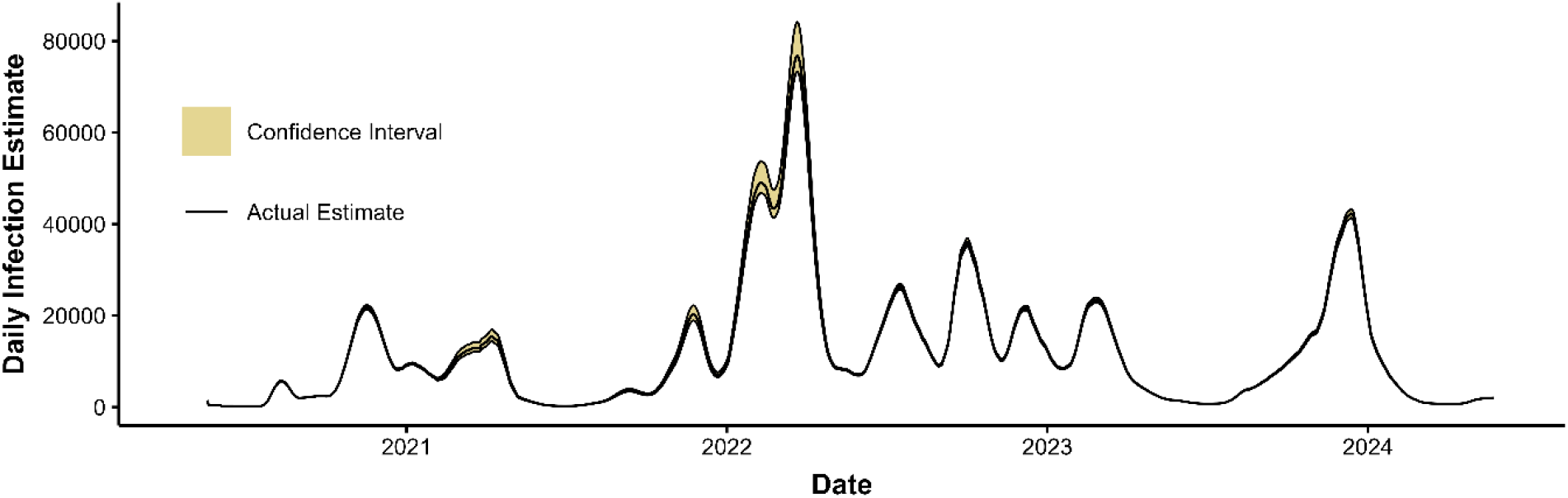
Daily infections estimated from wastewater data including lower and upper bounds as Confidence Intervals.

The wastewater model follows the trend of documented infections (7 day average) even in timeframes it was not previously applied to (Figure S5). The correlation between these two is r = .985 indicating perfectly matching trends.

**Figure S5:**
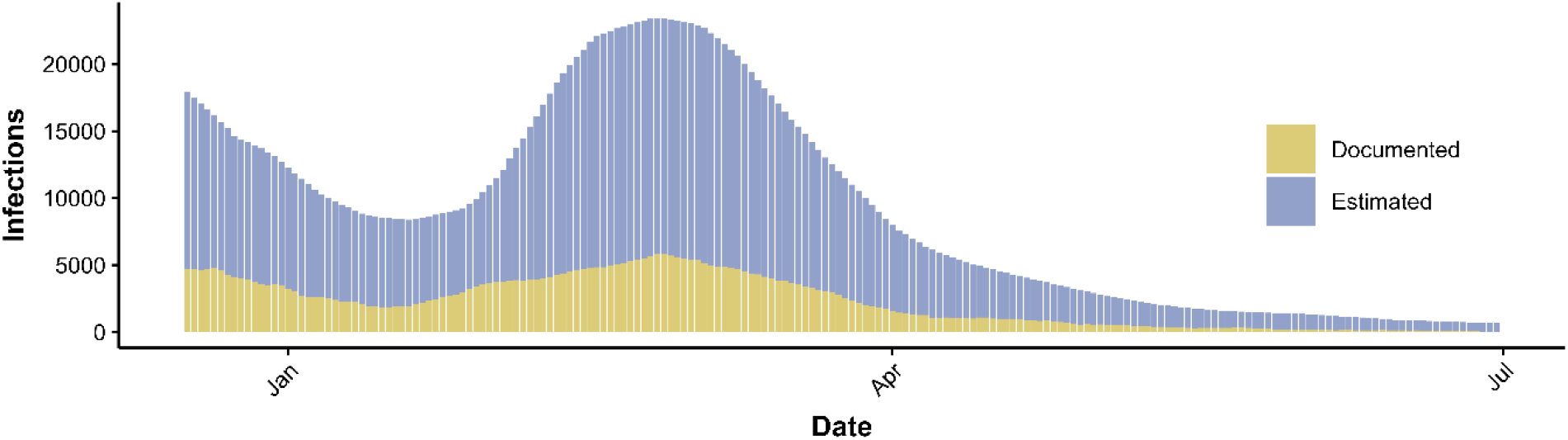
Documented and estimated infections between December 17, 2022 and June 30, 2023. Documented infections were averaged over 7 days.

##### Agent-based model

**Figure S6:**
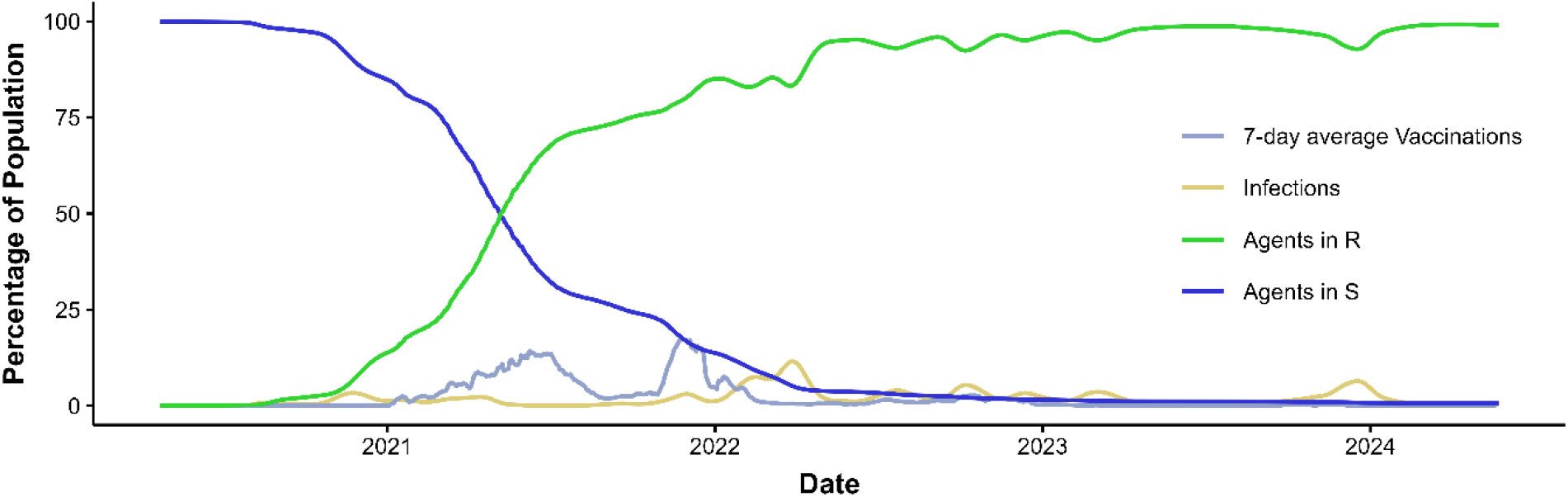
Time series of S, R, daily infections and daily vaccinations.

**Figure S7:**
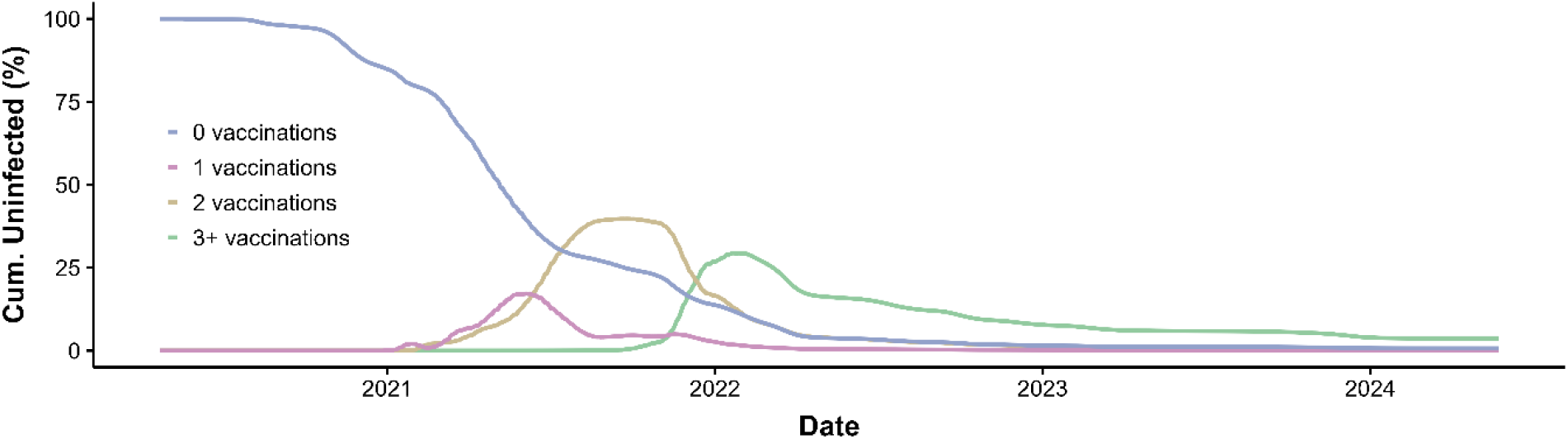
Percentage of people with no infection, stratified by number of vaccinations, by day.

**Figure S8:**
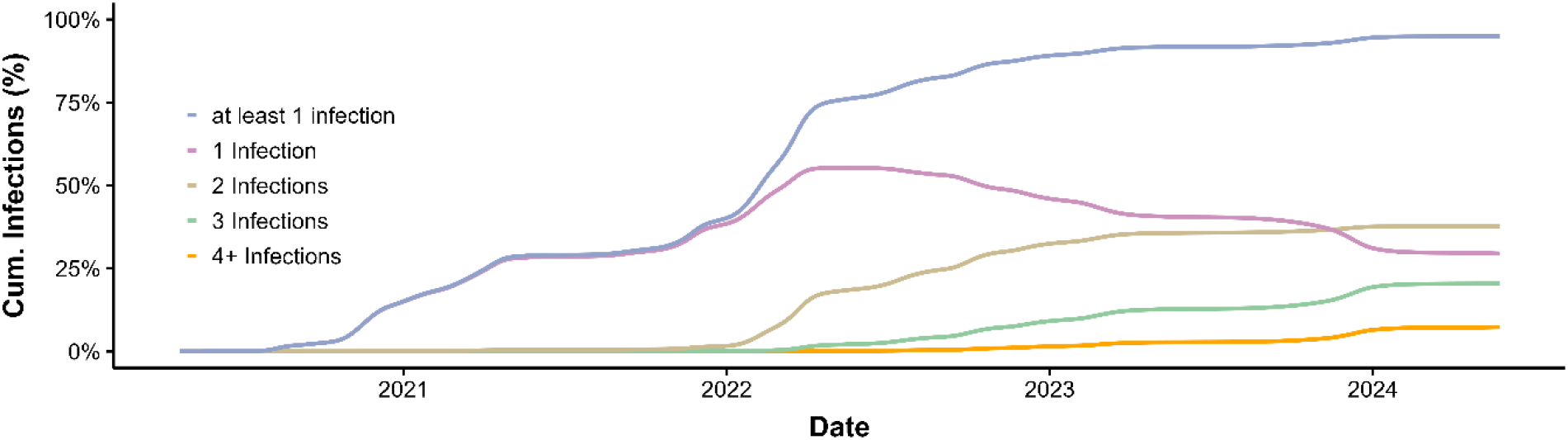
Percentage of people with at least one infection and stratified by number of infections, by day.

**Figure S9:**
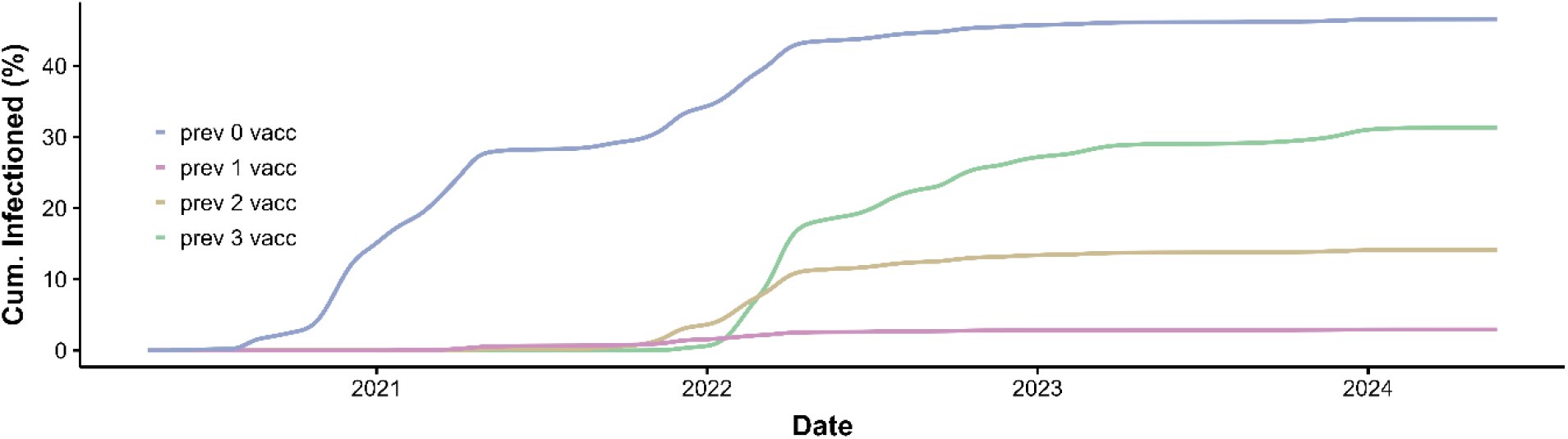
Cumulative percentage of population, stratified by vaccination status on day of first infection.

##### IFR Regression

**Figure S10:**
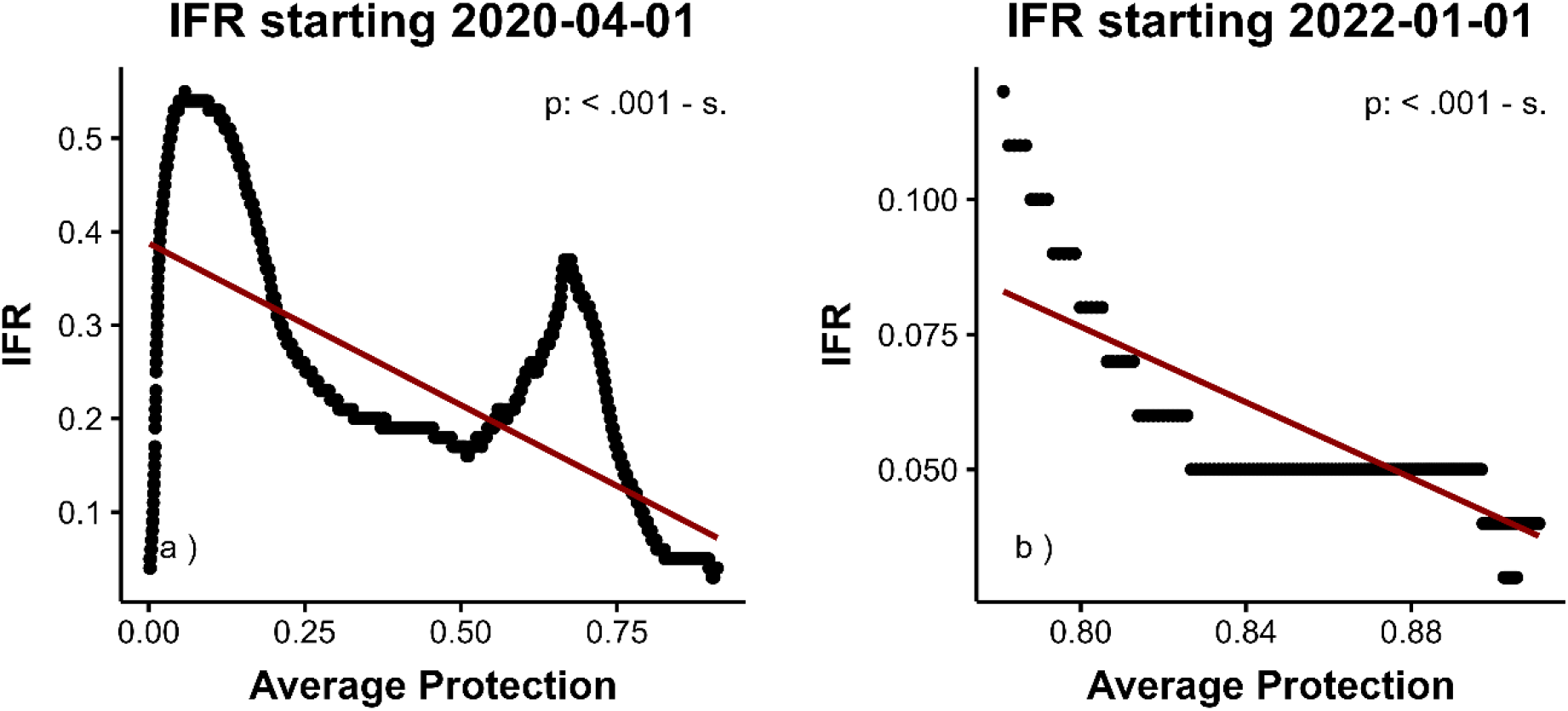
Regression lines of 4 month rolling window IFR on 4 month rolling window DP.

##### Sensitivity Analyses

**Table S3:**
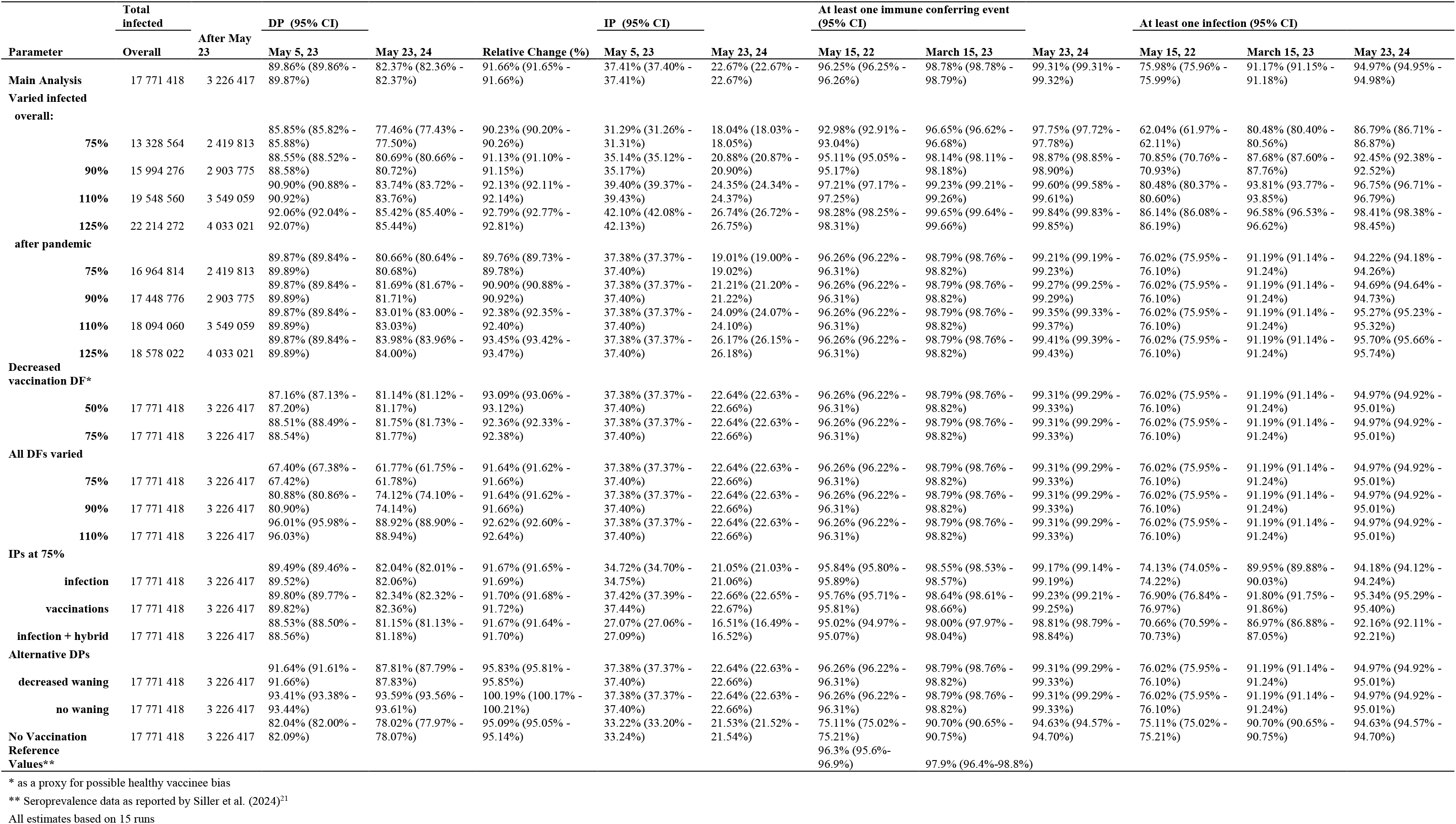
Different outcome values based on parameter choice.

##### Sensitivity Figures

**Figure S11:**
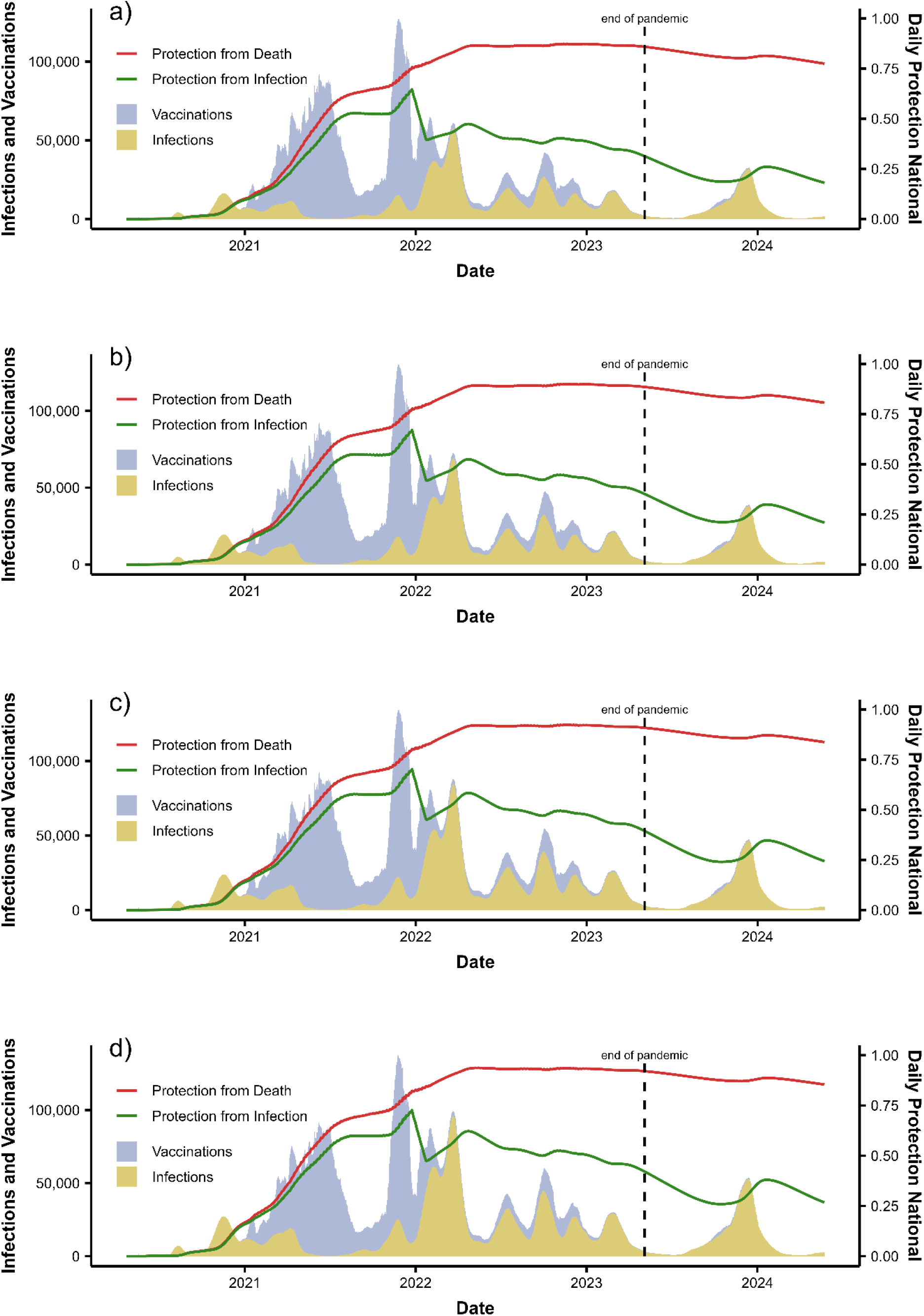
Changed overall daily infections, to (a) 75%, (b) 90%, (c) 110% and (d) 125% of the original estimate.

**Figure S12:**
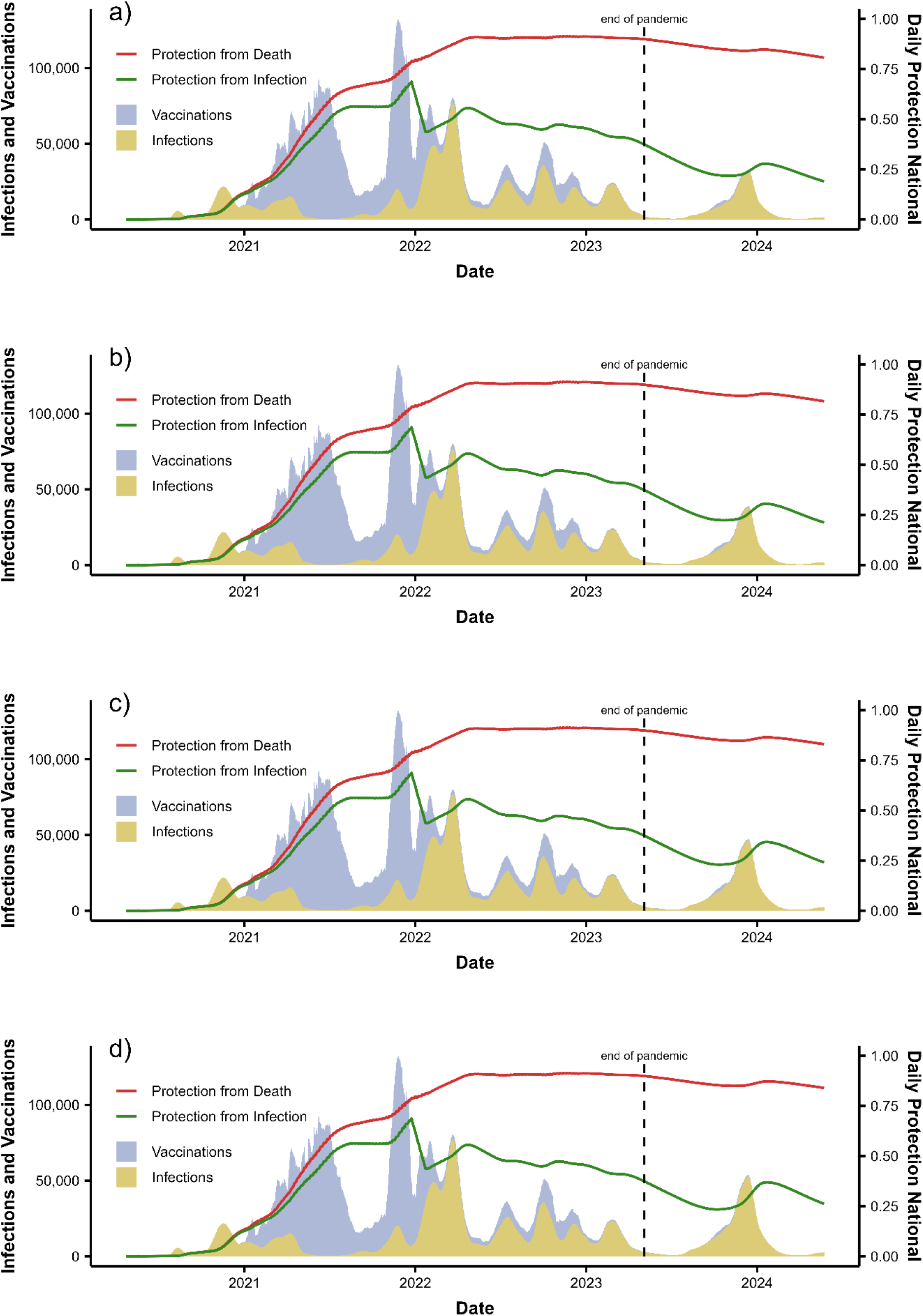
Changed daily infections after May 6, 2023, to (a) 75%, (b) 90%, (c) 110% and (d) 125% of the original estimate.

**Figure S13:**
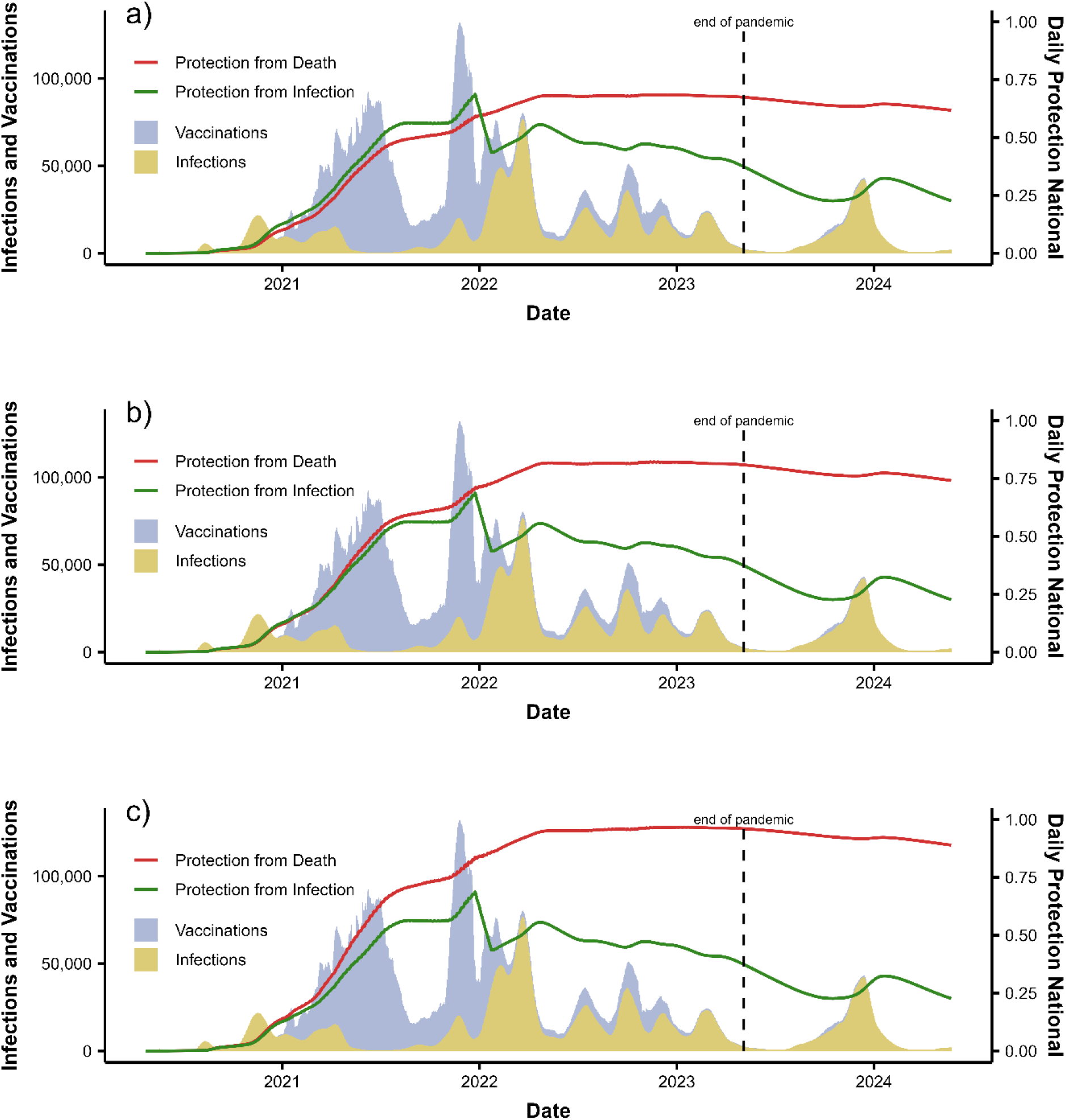
Varied all DP to (a) 75%, (b) 90% and (c) 110% of the original estimate.

**Figure S14:**
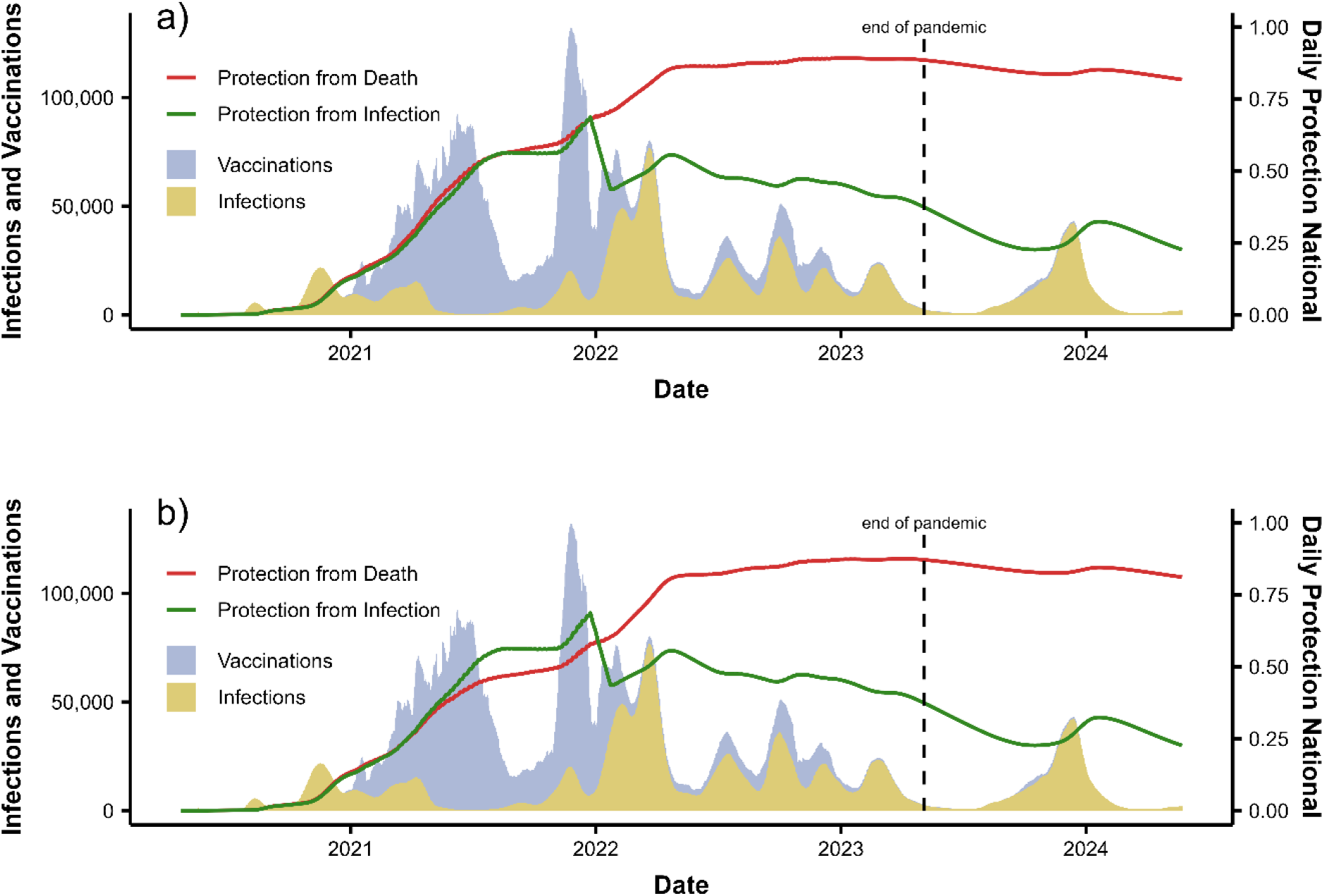
Reduced vaccination DP to (a) 75% and (b) 50% of the original estimate.

**Figure S15:**
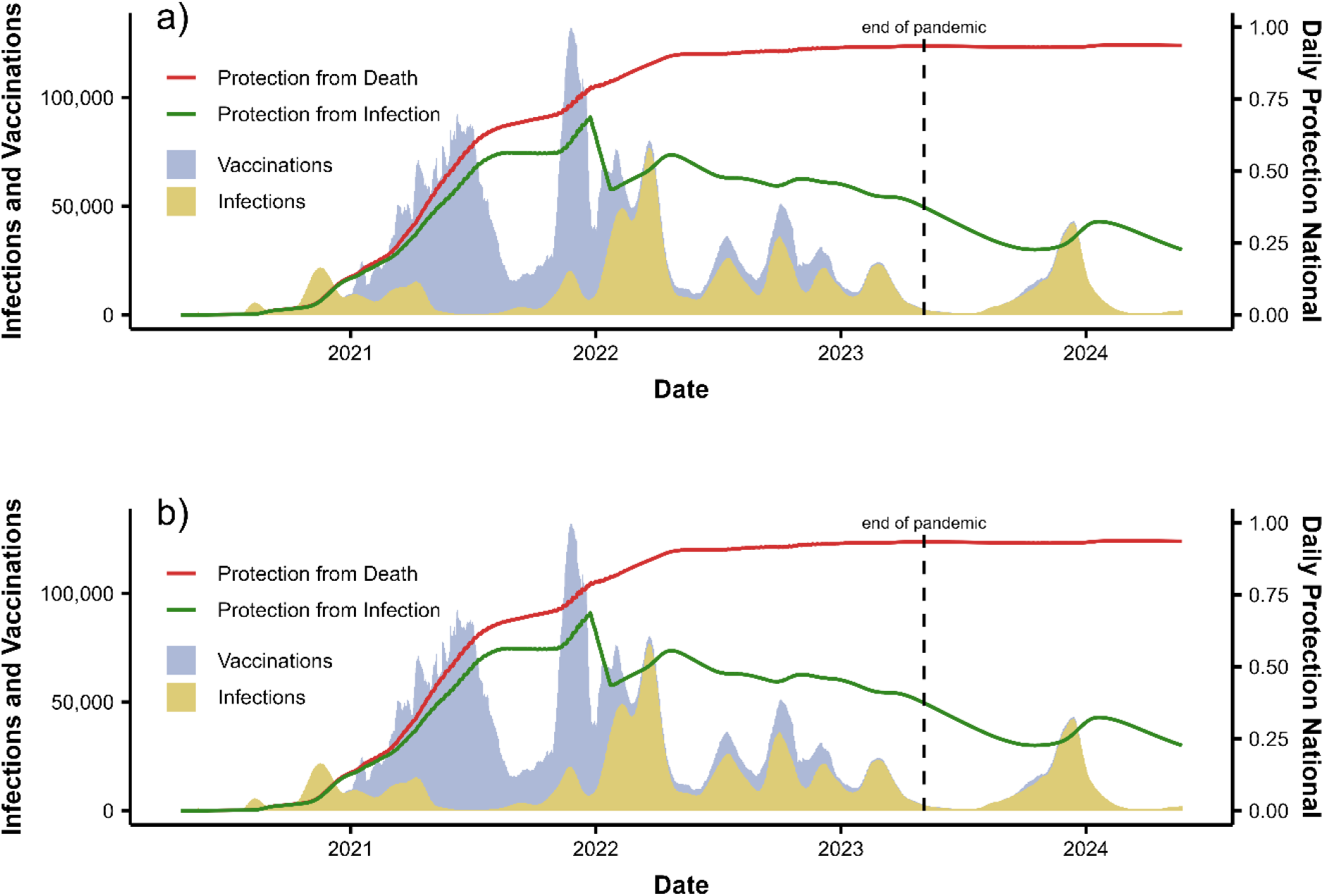
Alternative DP estimates. (a) DP mean of the main estimates and estimates without waning. (b) DP estimates without waning.

**Figure S16:**
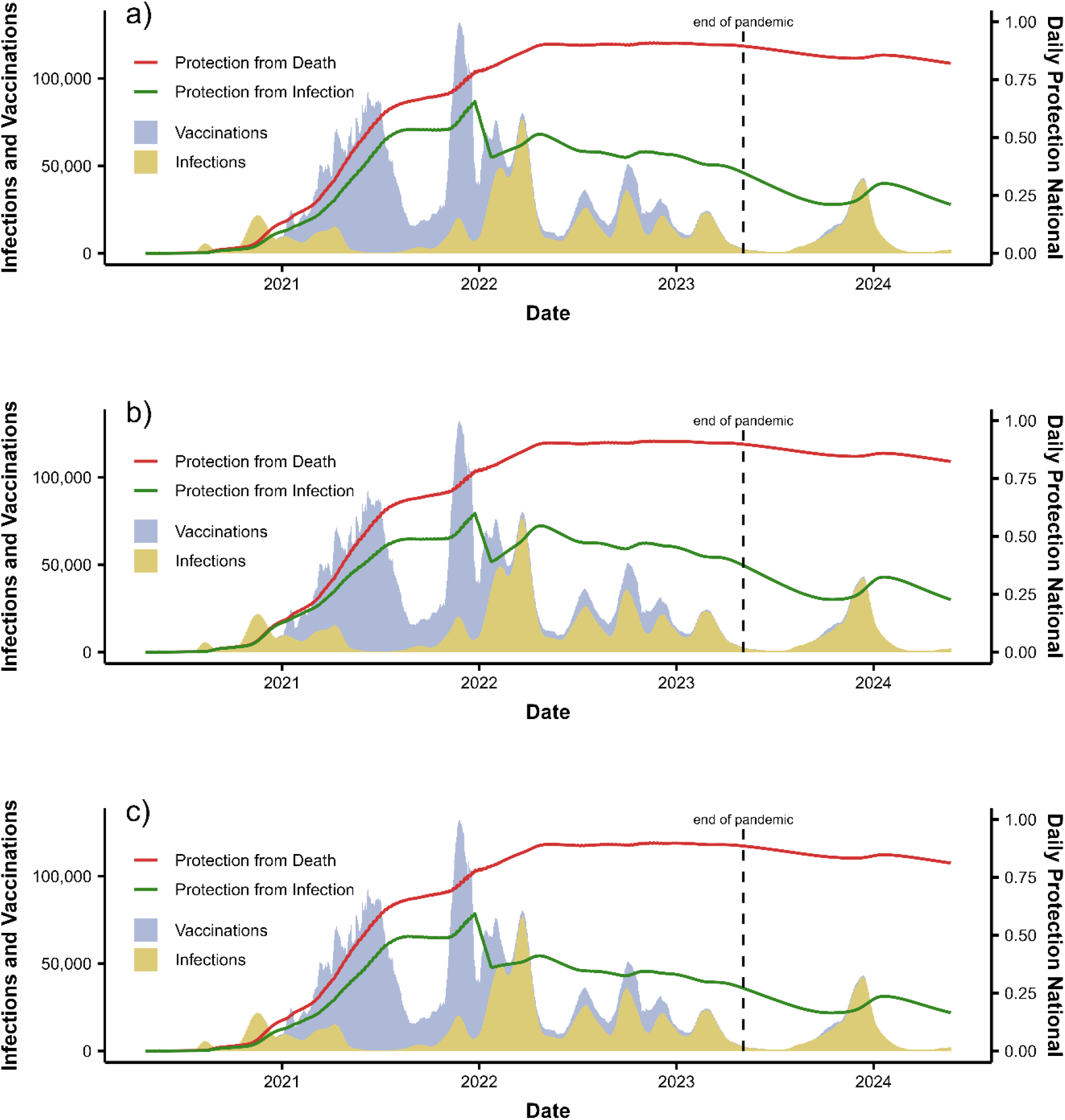
Reduced IP to 75% for different conditions. (a) IP of previous infection. (b) IP of all vaccination conditions. (c) IP of previous infection and hybrid immunity.

### Supplementary Discussion

#### Limitations

##### Wastewater

There are measurement errors in the determination of the hydrochemical parameter concentration. While outliers are addressed as mentioned in the pre-processing steps, smaller errors, akin to noise, cannot be avoid.

The shedding rate varies from person to person. The average shedding rate is also not constant. The immunization rate and the virus variant are potential factors that can also influence the average shedding rate.

##### Agent Based Model

We did not account for time between vaccinations. While there were suggestions set on time between first second and third vaccination, many early cases were rushed on purpose for high-risk groups. This could be addressed using a probability distribution based on the time since last vaccination, but we deemed the added complexity and runtime not worth the likely miniscule (if any) changes in values.

